# Effect of mHealth interventions on psychological issues experienced by women diagnosed with breast cancer receiving chemotherapy: A systematic review and meta-analysis

**DOI:** 10.1101/2022.02.17.22271140

**Authors:** Oluwadamilare Akingbade, Khanh Thi Nguyen, Ka Ming Chow

## Abstract

**Background:** A range of psychological issues often accompany breast cancer chemotherapy. Owing to their ubiquity, mobile phones have been used to deliver supportive interventions that address these issues. However, we currently lack sufficient evidence to guide the design of such interventions.

**Aim:** To analyse and synthesise available evidence on the effectiveness of mobile-phone-based (mHealth) interventions in alleviating the psychological issues experienced by women receiving chemotherapy for breast cancer.

**Methods:** A systematic literature search was conducted from 14 relevant databases. Revman 5.4 was used to pool the quantitative results from comparable studies for statistical meta-analysis. For clinically heterogeneous studies where statistical pooling of results was not possible, a narrative summary was used to present the findings.

**Results:** The review included ten published studies which covered 1,548 patients. The meta-analysis results indicated a significant improvement in the quality of life (standardised mean difference [SMD] = 0.32, 95% confidence interval [CI] [0.07, 0.58], *p* = .01, I^2^ = 17%). No significant effects were found for anxiety (SMD = -0.01, 95% CI [-0.26, 0.25], *p* = 0.96, I^2^ = 53%) and depression (SMD = 0.02, 95% CI [-0.17, 0.20], *p* = .87, I^2^ = 0%). Individual studies suggest reduced symptom prevalence (*p* = .033, *d* =.27), symptom distress (*p* = .004, *d =* .004), symptom interference (*p* = .02, *d =* .51), supportive care needs (*p* < .05, *d =* 2.43); improved self-efficacy (*p* = .03, *d =* 0.53), self-esteem (*p* <.001, *d* = 0.87) and emotional functioning (*p* = .008, *d =* .30). The methodological quality ranged from low to moderate.

**Conclusion:** mHealth interventions might be helpful in addressing certain psychological issues experienced by this population, although the evidence is still being gathered and not yet conclusive. More rigorous trials are hereby warranted to confirm the suitable duration while addressing the methodological flaws found in previous studies

**PROSPERO registration number:** CRD42021224307

## 1. Introduction

In 2020, one in eight cancers diagnosed worldwide was a type of breast cancer (BC); likewise, one in four of all new cancer cases in women was some form of BC [1]. In the same year, across the globe, female BC surpassed lung cancer to become the most prevalent cancer; BC affects 2.1 million women each year, with the number of new cases expected to increase by 46.5% by the year 2040 [1,2].

In addition to the high mortality rate of BC, the illness trajectory of BC patients can be extremely challenging. It has been estimated that approximately 50% of women diagnosed with BC will experience psychological issues at some point in their illness [3,4]. Psychological issues reported by this population include anxiety, depression, symptom distress, symptom prevalence, unmet supportive care needs, poor symptom control, increased symptom interference and reduced emotional functioning, all of which can reduce the quality of life [3-7].

Although psychological issues are common in cancer patients, they are not inevitable as appropriate interventions can mitigate the effects of these issues [7-10,11]. Evidence supporting the effectiveness of interventions in addressing the psychological issues experienced by women suffering from BC is being gathered. A systematic review and meta-analysis of the psychological interventions used for women with BC found that the interventions could be classified into three major categories: cognitive-behavioural therapy (CBT), psychoeducational therapy (PET) and supportive-expressive therapy (SET). They found a medium effect size of these interventions on anxiety, depression, mood, and the quality of life, especially for CBT and PET [12].

mHealth, defined as ‘health-related services delivered through mobile communication devices’ [13] has been recommended as a medium to deliver these interventions owing to the ubiquity of mobile phones [14]. As of 2009, more than 90% of the global population was documented to have access to mobile phone technology, with over two-thirds of the population owning a mobile phone [15]. In 2020, over three billion people worldwide had smartphones; this number is expected to grow by several 100 million in the years to come [16]. Mobile phone use is on the rise in low- and middle-income countries (LIMCs), as reported by the International Telecommunication Union (ITU) in 2015 [15]; they found that 70% of the seven billion mobile subscriptions across the world came from these countries. The WHO has also reported that mobile phones were more accessible than clean water in LIMCs [17].

As the popularity of mobile phones is growing, so is the body of evidence supporting their use in improving various medical conditions. The WHO has advocated for the application of mHealth interventions to manage chronic conditions like cancer, diabetes, and cardiovascular and respiratory diseases [17]. mHealth applications (apps) have proven useful in various fields such as pain management [18], obesity surgery [19] and diabetes [20]. They have been shown to augment the self-management of chronic diseases [21] and reduce the number of visits to healthcare centres, thereby reducing time wastage, distance travelled and the cost of accessing healthcare services [22] while simultaneously improving access to health information and communication between patients and healthcare teams [23].

Various randomised controlled trials (RCTs) have found mHealth interventions to be effective along the BC illness trajectory. For instance, mHealth interventions increased the number of women who underwent BC screening [24-27]. Similarly, in women who completed BC treatment, mHealth interventions provided effective psychosocial support, improving the quality of life and decreasing stress [28-30]. Furthermore, web-based exercise interventions were found to successfully increase the number of women who performed exercise [31-32]. Among women receiving treatment for BC, Jongerius and colleagues (2019) found that mHealth interventions were effective in promoting weight loss, decreasing stress and improving the quality of life [30]. Another study [33] also predicted that after the COVID-19 pandemic, telemedicine would be integrated into the care of oncology patients; however, sufficient evidence to guide this integration has not yet been established.

Although some systematic reviews have addressed the effects of telephone-based interventions on the care of women with BC [30,34-36], none of them has specifically assessed the effects on women undergoing chemotherapy. A meta-analysis of 20 telehealth interventions delivered at different time points along the BC trajectory found that the interventions improved the quality of life and self-efficacy and reduced depression, distress and perceived stress in the patients. However, the intervention did not have any significant effects on anxiety [34]. Similarly, another meta-analysis of 14 telephone-based interventions applied at different time points along the BC trajectory and found that the interventions reduced anxiety and improved the quality of life, but did not have any significant effects on depression [35]. Also, another review of research-tested applications delivered at different time points along the BC trajectory and found conflicting and inconclusive data regarding the effects of the interventions on psychological issues experienced by the patients [30]. Furthermore, a review of mobile applications (apps) used during the treatment of breast cancer revealed that the apps could be helpful in promoting self-care, reporting symptoms and adverse treatment-related effects; however, it was concluded that the current level of evidence is limited and the utility of the apps in women undergoing treatment for BC is still uncertain [36].

Given the limited evidence available for the effectiveness of mHealth interventions in women undergoing BC chemotherapy and to the best of our knowledge, no study has specifically addressed the effect of mHealth interventions on psychological issues in this population, a systematic review in this regard is warranted. A systematic review that specifically targets women undergoing chemotherapy for BC is required as the needs of these women differ from those of women in other stages of the BC trajectory, such as those who have completed treatment. Symptom clusters of pain, fatigue, anxiety and depression have been reported in women undergoing chemotherapy for BC [37-39], highlighting the need for interventions to address issues that specifically arise during chemotherapy. Although some mHealth interventions have been used for women with BC undergoing chemotherapy, the effects of these interventions on psychological issues remain unclear [30,40].

This review aims to identify, analyse, and synthesise the available evidence on mHealth interventions for addressing psychological issues experienced by women with BC undergoing chemotherapy. The objectives are to (1) identify the various mHealth interventions used for women with BC undergoing chemotherapy; (2) identify the effective components of these interventions; and (3) identify the effects of these interventions on psychological outcomes in this population.

## 2. Methods

We followed the Preferred Reporting Items for Systematic Reviews and Meta-analyses (PRISMA) guidelines [41]. The protocol of this systematic review was registered with the International Prospective Register of Systematic Reviews (PROSPERO registration number: CRD42021224307).

### 2.1. Search Strategy

The search strategies were formulated based on the characteristics of the databases (Appendix A, Table 1); our searches targeted articles that were available from the inception of the databases until January 2022. Fourteen databases were searched (Cochrane Library, British Nursing Index, Ovid Emcare, Allied and Complementary Medicine, Embase, Joanna Briggs Institute, Ovid Medline, Ovid Nursing, APA PsycINFO, Health and Medical Collections, Web of Science, Scopus, CINAHL and PubMed). The reference lists of the articles were also screened.

Titles, abstracts, keywords, and MeSH terms were used in the search. No date restrictions were applied. The key terms we used are listed in Appendix A.

### 2.2. Inclusion and Exclusion Criteria

Trials that focused on mHealth interventions to address psychological issues experienced by women with BC undergoing chemotherapy were included. Conference abstracts, protocols and ongoing studies were excluded. The details of the inclusion and exclusion criteria are described below.

#### 2.2.1. Population

Women (18 years and above), clinically diagnosed with BC and undergoing chemotherapy. Studies that included those who had completed treatment were excluded, as were studies that included women with BC but not undergoing chemotherapy.

##### Interventions

Trials that had assessed mHealth interventions delivered through any of the following media: telephone call and/or text; mobile phone support application; or internet-based support group. Interventions that were not delivered through mobile phones were excluded.

#### 2.2.2. Comparison

Participants in the comparison group should have received standard or usual care; participants receiving different kinds of interventions should have been compared with those receiving a mHealth intervention in the intervention group.

#### 2.2.3. Outcomes of interest

All studies that addressed psychological issues in women undergoing breast cancer chemotherapy were included in this review. Outcomes of interest were anxiety, depression, quality of life, supportive care needs, symptom burden, self-efficacy, social support, or self-esteem.

#### 2.2.4. Study design

Most of the included studies were RCTs. RCTs provide the strongest evidence for evaluating intervention effects [42]. One of the studies was a non-randomized controlled trial.

### 2.3. Study Selection

After the studies were identified from the databases, duplicates were removed with the aid of Refworks. The remaining articles were screened independently by two reviewers (AO and KNT, who are PhD students). Whenever the two reviewers failed to agree, a third reviewer (KMC) was consulted. Full texts of the potentially eligible articles were downloaded and assessed for eligibility (Figure 1).

**Figure 1:**
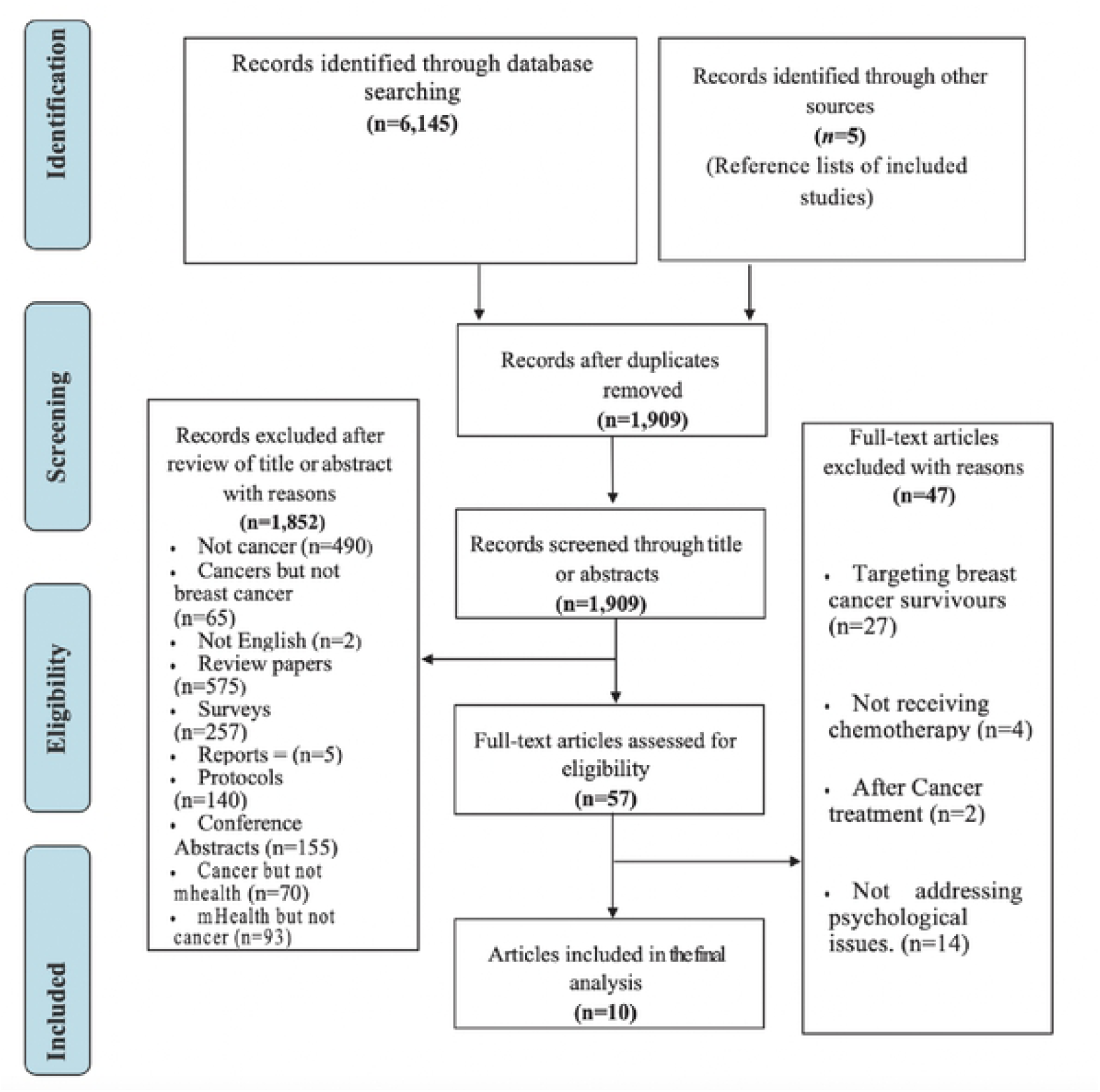
Study retrieval and selection flow chart (PRISMA)

### 2.4. Data Extraction

Data were extracted using a modified version of the Joanna Briggs Institute data extraction form [43]. Two reviewers independently conducted the data extraction and summarisation. Whenever an agreement could not be reached, a discussion was conducted between the two reviewers to resolve the differences. However, the opinion of a third reviewer was sought when a consensus could not be reached. The extracted data were population and participant demographics; settings; participant inclusion and exclusion criteria; content and duration of the intervention; providers; study methods, formats, outcome measures and results.

### 2.5. Methodological Quality

The methodological quality was assessed using the Joanna Briggs Institute (JBI) Critical Appraisal Checklist [44]. Thirteen questions were featured in the checklist and covered areas such as true randomisation, allocation concealment, blinding, outcome assessment, loss to follow-up, intention-to-treat analysis, statistical analysis, and trial design. Each question could be answered with ‘Yes’, ‘No’ or ‘Some concerns. Disagreements were resolved through discussions between the two reviewers.

### 2.6. Data Analysis

Revman 5.4 was used to pool the quantitative results of comparable studies for statistical meta-analysis. As the studies reported continuous data and the measuring scales for the outcomes were not identical for all the studies, we calculated the standardised mean differences (SMDs) and their 95% confidence intervals (CIs). The clinical heterogeneity of the studies was assessed by considering the similarities in the participants’ characteristics, study settings and designs, details of the interventions and outcome measures. The statistical heterogeneity of the studies was examined using the I^2^ test, with values of 0–30% being classified as low, 31–60% as moderate and 61–100% as high and data were pooled using random-effects model [45]. For clinically heterogeneous studies for which statistical pooling of results was not possible, a narrative summary was used to present the findings.

#### 2.6.1 Sensitivity analysis

Sensitivity analysis was conducted with the Revman 5.4 software by excluding one trial at a time based on the methodological quality as recommended by the JBI manual for evidence synthesis [46]. Also, the impact of altering between random and fixed effect models was explored. This was done to detect if any study was particularly influential in altering the results.

## Results

### 2.7. Search Results

The database search yielded 6,145 articles, alongside 5 articles that were retrieved through a manual search. Of these, 1,909 articles remained after removing the duplicates. After examining the titles and abstracts, 1,852 articles were excluded as they did not meet the inclusion criteria. The full texts of the remaining 57 articles were then reviewed, of which 47 were further excluded. Finally, 10 studies were selected for the analysis. The study retrieval and selection process are presented in Figure 1.

### 2.8. Study Characteristics

The 10 included studies were conducted in nine countries: Sweden [47], Japan [48], Taiwan [49], Iran [50, 54], South Korea [51], the United States [52], China [53], Canada [55], and Slovenia [56]. All the studies included a total of 1,548 women with BC undergoing chemotherapy. The studies were conducted between 2014 and 2021, with sample sizes ranging from 62 to 580.

### 2.9. Study Population

Five of the studies recruited only women with BC undergoing chemotherapy [47-49, 53, 55], the remaining five recruited those receiving other treatments (radiotherapy, surgery, and hormonal therapy) alongside chemotherapy [50-52,54,56].

### 2.10. Interventions

#### 2.10.1. Interveners

Nurses were the interveners in six studies [47-50, 54, 55], whereas in other studies the interventions were delivered by both nurses and doctors [53], a medical doctor [50], graduate-level health professionals [51] and oncologist [56].

#### 2.10.2. Characteristics of interventions

Regarding the delivery mode, the interventions in seven studies were app-based [46-48, 50, 53, 54, 56], one intervention was delivered through an Internet support group [51] and two studies through telephone calls [49,55]. With respect to duration, three studies [47,48,53] administered interventions for 12 weeks, one for three weeks [50], two for four weeks [50, 54], one for six weeks [52], one for 18 weeks [47], one from the beginning of the chemotherapy cycle to 8-10 days after the start of each cycle [55], and one spanned from their first week of treatment to the entire treatment period [56].

We identified four major components of interventions. All except two [43,56] of the studies had a component for BC education; five had a component for expert consultation [48-50, 53, 54], four had a component for group interaction [49,52-54], and five had a component for the self-reporting of symptoms [47-49, 55, 56]. One intervention also had a game-based learning component [51].

With respect to the time frame, the interventions could be accessed either anytime during the day [48,49,53,54]; from 8 am to 4 pm daily [47]; for 90 minutes daily [52]; for more than 30 minutes daily [51]; or through a 15–20 minute-long call twice a week [50]; or through two structured follow-up calls 24-72 hours and 8-10 days after the start of each cycle [55]; and one through the entire treatment period [56].

Regarding the frequency, the interventions were delivered either daily [47,48,52-54]; five days a week [47]; twice a week [50] or three times a week [51]; and others through the treatment period [55, 56].

All the studies had two-group designs. Only one study administered interventions to both groups, in which a standard internet support group was compared with a pro-social internet support group [52]. All the included studies conducted a pre-test and post-test to evaluate the outcomes. Five of the studies had three-time points for data collection [49,50,53,55, 56], four studies had two-time points [47,51,52, 54] and one study had four-time points [48]. The attrition rate ranged from 3% to 13% across the studies. Only one of the interventions had a theoretical underpinning, which was Bandura’s self-efficacy and social exchange theories [53]. The details of the interventions are listed in Appendix B.

### 2.11. Methodological Quality of Included Studies

All the studies reliably measured outcomes in the same manner for the treatment groups and followed the appropriate statistical analyses and trial designs. The groups were similar at baseline for all the studies. However, loss to follow-up was not adequately addressed in five of the studies, although the intention-to-treat analysis pointed to attrition bias [48,50,51,54,56]. None of the studies blinded the intervention providers as they were fully involved in the process and blinding was not feasible. The outcome assessors were not blinded in eight of the studies [47-51, 54-56]. Allocation concealment was not done in seven of the studies [48-51, 54-56]. Only one of the studies blinded the participants [49]. Overall, the studies had low to moderate methodological quality. (Appendix C).

### 3.6 Result of Sensitivity Analysis

Sensitivity analysis done to detect the undue influence of any trial did not reveal the presence of such occurrence. Similarly, altering between fixed and random effects models did not materially alter the results.

### 3.7 Outcomes

Anxiety was measured in seven studies [48,51-56] and depression was measured in six studies [48,51-53, 55-56], quality of life was assessed in six of the studies [47,49,51,53, 55-56], self-efficacy was measured in two studies [53, 55], symptom distress [47], supportive care needs [50], and self-esteem [54] were also evaluated in one study each. Sixteen instruments were used to measure outcomes across all the studies (Appendix B).

### 3.8 Effects of mHealth Interventions on Psychological Issues

#### 3.8.1 Quality of life

Three comparable studies that measured the effects of mHealth intervention on quality of life in 286 participants were pooled and included in the meta-analysis [49,51,53]. The quality of life was improved significantly after completion of the interventions (SMD = 0.32, 95% CI [0.07, 0.58], *p* = .01, I^2^=17%) (Figure 2).

**Figure 2:**
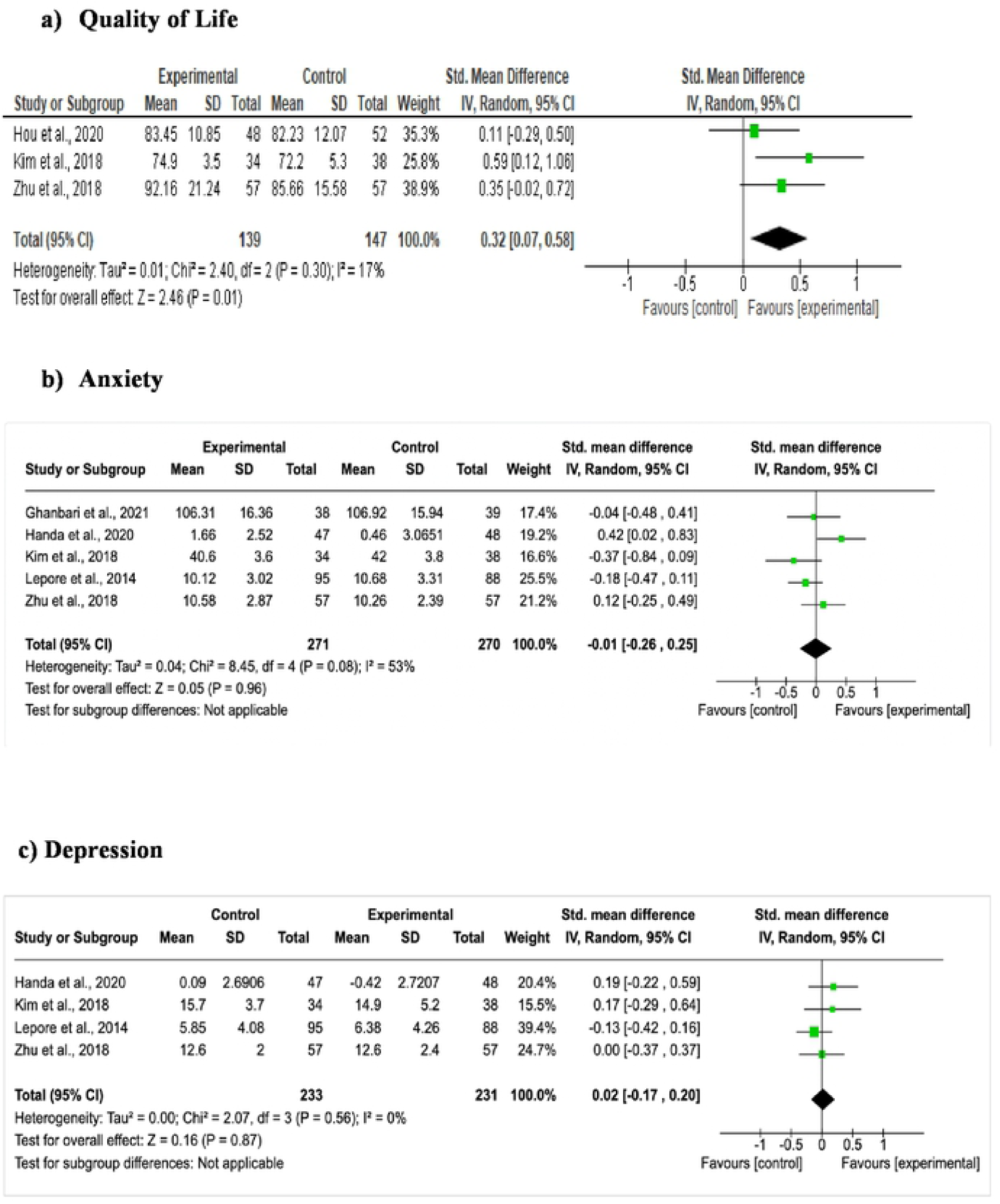
Forest plots-Effects of mhealth interventions on quality of life, anxiety and depression.

#### 3.8.2 Anxiety and depression

Five comparable studies that measured anxiety in 541 participants [48,51-54] and four comparable studies that measured depression in 464 participants [48,51-53] were pooled and included in the meta-analysis. After completion of the intervention, no significant effects were found for anxiety (SMD = -0.01, 95% CI [-0.26, 0.25], *p* = 0.96, I^2^=53%) and depression (SMD = 0.02, 95% CI [-0.17, 0.20]; *p* = .87, I^2^=0%) (Figure 2).

#### 3.8.3 Symptom prevalence, distress, and interference

One study [47] measured symptom prevalence and distress. The intervention group reported significant reductions in symptom prevalence (*p* = .033, *d* =.27), nausea (*p* = .041), vomiting (*p* = .037) and sadness (*p* = .003). The overall symptom distress (*p* = .004, *d =* .004), and physical symptom distress (*p* = .031) were also found to be significantly lower in the intervention group. Emotional functioning was found to be significantly higher in the intervention group (*p* = .008, *d =* .30). One study measured symptom interference and found it to be lower in the intervention group (*p* = .02, *d =* .51) [53].

#### 3.8.4 Supportive care needs

One study [50] tested the effectiveness of a telephone-based support system and found that the mean score for supportive care needs in the intervention group was significantly lower than in the control group supportive care needs (*p* < .05, *d =* 2.43).

#### 3.8.5 Self- efficacy

One study [53] compared the effectiveness of an app named *BC e-Support Programme* with that of the usual care regime. They found significant improvements in self-efficacy using the app (*p* = .03), but this effect was not sustained after 6 months. However, another study that compared the effectiveness of telephone-based management for patients receiving chemotherapy found no significant improvement in self-efficacy [55].

#### 3.8.6 Self-esteem

One study [54] that compared the effects of psychoeducational interventions using a mobile app with that of the usual care found a significant improvement in self-esteem (*p* <.001, *d*=0.87).

## 4.0 Discussion

Given the wide use of mobile phones and their increasing applications in breast oncology, a systematic review was conducted to evaluate the effectiveness of miHealth interventions on alleviating the psychological issues experienced by women with BC undergoing chemotherapy. To the best of our knowledge, this is the first systematic review and meta-analysis addressing the usage of mHealth interventions in managing psychological issues in this population. This review was conducted in a bid to provide evidence for the design of mHealth interventions for women diagnosed with BC and undergoing chemotherapy. Such evidence has not yet been collated, even though the application of mHealth interventions at different time points along the BC illness trajectory has gained prominence. The needs of women differ as they progress along their BC illness trajectories; for instance, the needs of those undergoing treatment are different from the needs of women who have completed treatment. Therefore, a systematic review focusing solely on those receiving chemotherapy was required as a number of psychological issues, such as anxiety, depression, symptom distress, poor symptom control, increased symptom interference, fear of death and unmet supportive care needs, have been reported in this population [3, 4, 6, 7].

The first objective of this review was to identify the different mHealth intervention types available for women with BC undergoing chemotherapy. This review shows that most of the interventions that were assessed were delivered through mobile apps [46-48, 50, 53, 54, 56]. This is not unexpected; a review by Mobasheri and colleagues [14] including BC mobile apps found that the number of such apps had increased since they were first developed in 2009. A total of 185 apps used for the delivery of mHealth interventions was found, although most of them were not evidence-based as they were not research-tested and lacked the involvement of health professionals [14]. However, Jongerius and colleagues [30] conducted a systematic review specifically on research-tested apps used for BC prevention and treatment and for women who completed treatment; they found 29 mobile apps, of which seven focused on BC prevention and early detection, 12 focused on care management and 10 were directed at women who had completed treatment for BC [30]. This review confirmed the increasing popularity of BC apps to deliver interventions at different points along the cancer trajectory. However, researchers are still gathering evidence on the usage of these apps.

The second objective of this review was to identify the effective components of these interventions. We found that in these app-based designs, four major components, i.e., BC education, group interaction, self-reporting of symptoms and expert consultation, were effective in addressing some of the psychological issues experienced by this population [47-50, 52-56]. Game-based learning was another component that was used in one study [51]. The intervention found no significant effect on mood and anxiety, although the quality of life improved with the intervention and there was better drug adherence. Although the use of educational games improved psychosocial functions such as self-esteem, self-efficacy and depression in paediatric patients with obesity [57], the effect of game-based learning on psychological issues of women diagnosed with BC undergoing chemotherapy is not yet confirmed. This highlights the need for more interventions.

We also observed that most of the interventions were delivered by nurses [47-50,53]. The interventions provided by nurses improved the quality of life, emotional functioning and self-efficacy and reduced the supportive care needs, symptom prevalence, symptom distress and symptom interference in patients. This echoes the findings of a systematic review of psychological care delivered to cancer patients over a 25-year period and found that most of the interventions were delivered by nurses, closely followed by psychologists [11]. Also, another review on the effects of psychoeducational interventions on gynaecological cancer patients and concluded that nurses were the ideal providers of psychoeducational interventions as they facilitated better outcomes following the interventions [58]. As most of the studies in this review were psychoeducational interventions [49-54], we infer that nurses have a pivotal role in providing psychological care for women with BC undergoing chemotherapy. As nurses spend more time with patients [59], providing direct patient care [60]; we might infer that they occupy a vantage position in rendering psychological support for patients while chemotherapy is ongoing. Indeed, a qualitative study of women with BC undergoing chemotherapy reported that patients verbalised the need for more support from nurses [61].

As the duration of the interventions varied from three weeks to 18 weeks, as did the outcomes of these different durations, we were unable to determine the most effective duration for interventions. Also, we were unable to identify the efficacy of the intervention components since most of the interventions featured multiple components. This highlights the need for more rigorous interventions and investigations to determine optimal durations.

The third objective of this review was to document the effectiveness of mHealth interventions in the population of women with BC undergoing chemotherapy. We found that mHealth interventions might be effective in addressing some psychological issues faced by this population. For the individual studies, we found that the interventions improved emotional functioning, self-efficacy, self-esteem, and reduced the supportive care needs, symptom prevalence, symptom distress and symptom interference. However, these studies are limited, and the evidence is still being gathered. For the meta-analytic findings, the effects of mHealth interventions were effective in improving quality of life. This was similar to the findings of two previous meta-analyses that also found a significant improvement in the quality of life [34,35]. However, the intervention effects on anxiety and depression remain unclear as there was no significant difference between the control and intervention groups on both outcomes. Two previous meta-analyses of the effects of mobile phone-based interventions on anxiety and depression in this population also yielded contrasting results [34,35]. One study found anxiety, but not depression, to be significantly reduced [35], whereas the other study reported the inverse [34]. These inconsistencies point to the need for further research to test the effectiveness of mHealth interventions on anxiety and depression as supported by two studies included in this review [49,53].

Some research gaps were found in this review. As the field is relatively new, with the earliest intervention included in this review having been conducted in 2014, the evidence is still being gathered, and more rigorous trials are warranted. Only one study used a theoretical framework to underpin the intervention [53] indicating the need for more theory-based mHealth interventions. Similarly, low to moderate heterogeneity was found among the interventions included in the meta-analysis. Furthermore, most of the studies were conducted in developed countries. This underscores the requirement for more studies in developing countries as in 2015, the ITU reported that 70% of the seven billion global mobile phone subscriptions came from LMICs [15].

## 4 Study Limitations

The review has some limitations. It is difficult to generalise our findings due to the heterogeneity of the studies. We also only considered trials published in English and excluded conference papers, theses, and grey literature. Moreover, the nascence of the field implies the existence of very few studies on the subject. We found methodological flaws such as a lack of blinding, allocation concealment and intention-to-treat analysis were found among some studies in this review. Therefore, these limitations must be considered when interpreting our findings.

## 5 Clinical Implications

The four major components, namely, BC education, group interaction, self-reporting of symptoms and expert consultation, identified in this review should be considered when designing future mHealth interventions. Furthermore, nurses, who constituted most intervention providers, are crucial for the delivery of mHealth interventions as their intervention significantly improved some psychological outcomes in this population. However, the optimal duration of the intervention is yet to be determined as the durations included in this review varied widely. Owing to the various methodological flaws found in the studies reviewed, there is a need to design more rigorous trials in which these flaws can be addressed. The development of mHealth interventions should be guided by appropriate theories to the target population. More studies are needed to determine the most effective intervention designs.

## 6 Conclusion

This systematic review focused on mHealth interventions in women with BC receiving chemotherapy. The review suggested that the interventions were effective in reducing certain psychological issues such as symptom prevalence, symptom distress, supportive care needs, and improving self-esteem, emotional functioning, and quality of life. However, the evidence in favour of these interventions is still being gathered and is not yet conclusive. Further rigorous studies are warranted to address the methodological flaws identified in this review. In conclusion, mHealth interventions might be effective in providing psychological support for women with BC receiving chemotherapy.

## Data Availability

All relevant data are within the manuscript and its Supporting Information files.

## 7 Declarations

### Availability of data and material

(data transparency): Not applicable

### Code availability

(software application or custom code): Not applicable

### Authors’ contributions

Conception of Idea: AO

Literature Search: AO

Data Analysis: AO, KNT, KMC

Draft: AO, KMC

Critical revision: KMC

### Ethics approval

(include appropriate approvals or waivers): Not applicable

### Consent to participate

(include appropriate statements): Not applicable

### Consent for publication

(include appropriate statements): Not applicable

